# Pain Catastrophizing, Pain Self-Efficacy, and their Interaction as Predictors of Health Outcomes in Chronic Pain

**DOI:** 10.64898/2026.06.15.26355697

**Authors:** Emma Raney, Troy C. Dildine, Samsuk Kim, Sean C. Mackey, Dokyoung S. You

**Affiliations:** Stanford School of Medicine, Department of Anesthesiology, Perioperative, and Pain Medicine, Stanford University, Palo Alto, CA USA; Dept. of Family and Community Medicine University of Oklahoma Health Sciences, Tulsa, OK, USA

**Keywords:** Pain Self-Efficacy, Pain Catastrophizing, Physical Function, Psychosocial Health Outcomes

## Abstract

**Introduction:** Pain catastrophizing and pain self-efficacy are well-established predictors of health outcomes in chronic pain. Higher pain catastrophizing, a maladaptive cognitive process, predicts worse health outcomes, whereas higher pain self-efficacy, an adaptive cognitive process, predicts better health outcomes. This study examined whether pain catastrophizing and pain self-efficacy interactions predict physical and psychosocial health outcomes at 3 months and their change over 3-months among patients with chronic pain who sought care at a tertiary pain clinic.

**Methods:** Adults with chronic pain (N = 181; 66.7% female; M_age_ = 58.7) completed baseline assessments of the Pain Catastrophizing Scale (PCS), Chronic Pain Self-Efficacy Scale (CPSS), and PROMIS measures of physical (pain intensity, pain interference, physical function) and psychosocial health (depression, anxiety, anger, loneliness). PROMIS measures were repeated at 3 months. Hierarchical multiple regression analyses tested PCS, CPSS, and their interaction as predictors of outcomes at 3 months and change scores from baseline to 3 months.

**Results:** The PCS×CPSS interaction significantly improved prediction for physical function (ΔR^2^ = 0.02, p = .02). Higher baseline self-efficacy predicted better physical function (β = 0.65, p < .001), but this effect weakened with higher levels of pain catastrophizing. The interaction also predicted change scores in physical function (p = .025) but was marginal after false discovery rate correction (p = .059). Additionally, a significant interaction emerged for loneliness change scores (p = .01): higher self-efficacy predicted greater reductions in loneliness, attenuated by higher catastrophizing.

**Conclusion:** Pain self-efficacy interacted with pain catastrophizing to predict physical function and loneliness at 3 months. Greater self-efficacy was associated with better outcomes, with associations diminished with higher levels of pain catastrophizing. Findings highlight the moderating role of adaptive and maladaptive cognitions and suggest interventions should address both processes to optimize recovery in physical and social functioning.

**Key Message:** An interaction between pain catastrophizing and pain self-efficacy predicted physical function and changes in loneliness at 3 months. Greater self-efficacy was associated with better physical function and greater decreases in loneliness, but these associations weakened as catastrophizing increased. These findings highlight how maladaptive and adaptive pain-related cognitions interacted to predict physical and social function in chronic pain.

## Introduction

Chronic pain affects an estimated 50.2 million adults in the United States and represents a substantial yet under-addressed public health challenge^1^. It is a leading cause of disability, work loss, and reduced quality of life, with significant financial burden^2^. Individuals living with chronic pain frequently report limitations in both physical and psychosocial functioning^3–4^. Several modifiable psychological factors have been identified as potential targets to mitigate these negative consequences, most notably pain catastrophizing and pain self-efficacy^5–6^. Pain catastrophizing refers to maladaptive cognitive and emotional responses to pain, characterized by magnification, rumination, and helplessness. It is one of the most extensively studied and validated constructs in pain research^7–8^ and consistently predicts greater pain intensity, disability, and psychological distress^9^. However, with large patient heterogeneity within chronic pain, other psychosocial factors (e.g., self-efficacy) are likely important for chronic pain outcomes.

Self-efficacy describes the process of individuals initiating cognitive resources to cope and persist despite obstacles and aversive experiences^10^. Pain self-efficacy narrows this definition to adaptive cognitive processes related to pain, i.e., the belief in one’s ability to function and manage pain effectively^11^. Chronic pain requires continuous adjustment and development of effective coping strategies to minimize its impact on daily functioning. Consistent with this theoretical framework, pain self-efficacy predicts physical function, disability, and emotional distress^12^. Despite its clinical relevance, pain self-efficacy is not commonly assessed in practice^13^ and small sample sizes; therefore, evidence is limited but suggest it predicts pain-related outcomes and depression, but its relation to other psychosocial outcomes (e.g., loneliness) is lacking.

In the context of chronic pain, individuals may experience both maladaptive and adaptive cognitive responses^14–15^. High self-efficacy may counteract the negative impact of catastrophizing on pain intensity, disability, and emotional distress, whereas high catastrophizing may weaken or negate the advantages conferred by strong self-efficacy. However, the extent to which pain catastrophizing and pain self-efficacy interact to influence later physical or psychosocial health outcomes remains unclear. Clarifying this interaction is important, as understanding how these factors interplay may better explain differences in patients’ adjustment and progress over time.

This study aimed to examine whether the interaction between pain catastrophizing and pain self-efficacy predicts (1) physical outcomes (pain intensity, pain interference, physical function) and psychosocial outcomes (depression, anxiety, anger, loneliness) at 3-month follow-up, and (2) changes in these outcomes from baseline to 3-months among adults with chronic pain. We hypothesized that: 1) the interaction between pain catastrophizing and pain self-efficacy would significantly predict physical and psychosocial outcomes at the 3-month follow-up, above and beyond the main effects and 2) the interaction between pain catastrophizing and pain self-efficacy would significantly predict changes in physical and psychosocial outcomes over time.

## Methods

### Sample

Stanford University’s Institutional Review Board approved all study procedures. Participants were recruited from the Stanford Pain Management Center using our Learning Health System, CHOIR^16^, between April and November 2023, with the last follow-up in January 2024. Adults (≥18) with chronic pain were eligible, and no exclusion criteria were applied. The primary study aimed to identify social determinants of high-impact chronic pain. The Chronic Pain Self-Efficacy Scale (CPSS^17^) was added mid-study (July 2023–January 2024). During this period, 256 participants completed informed consent and baseline surveys via REDCap^18^, a HIPAA-compliant database. At 3 months, 185 participants (72.3%) completed follow-up; 4 participants’ missing age data were excluded, yielding a final sample of 181. Baseline demographic characteristics and patient-reported outcomes did not significantly differ between those who were included and excluded participants (see Table 1).

**Table 1.**
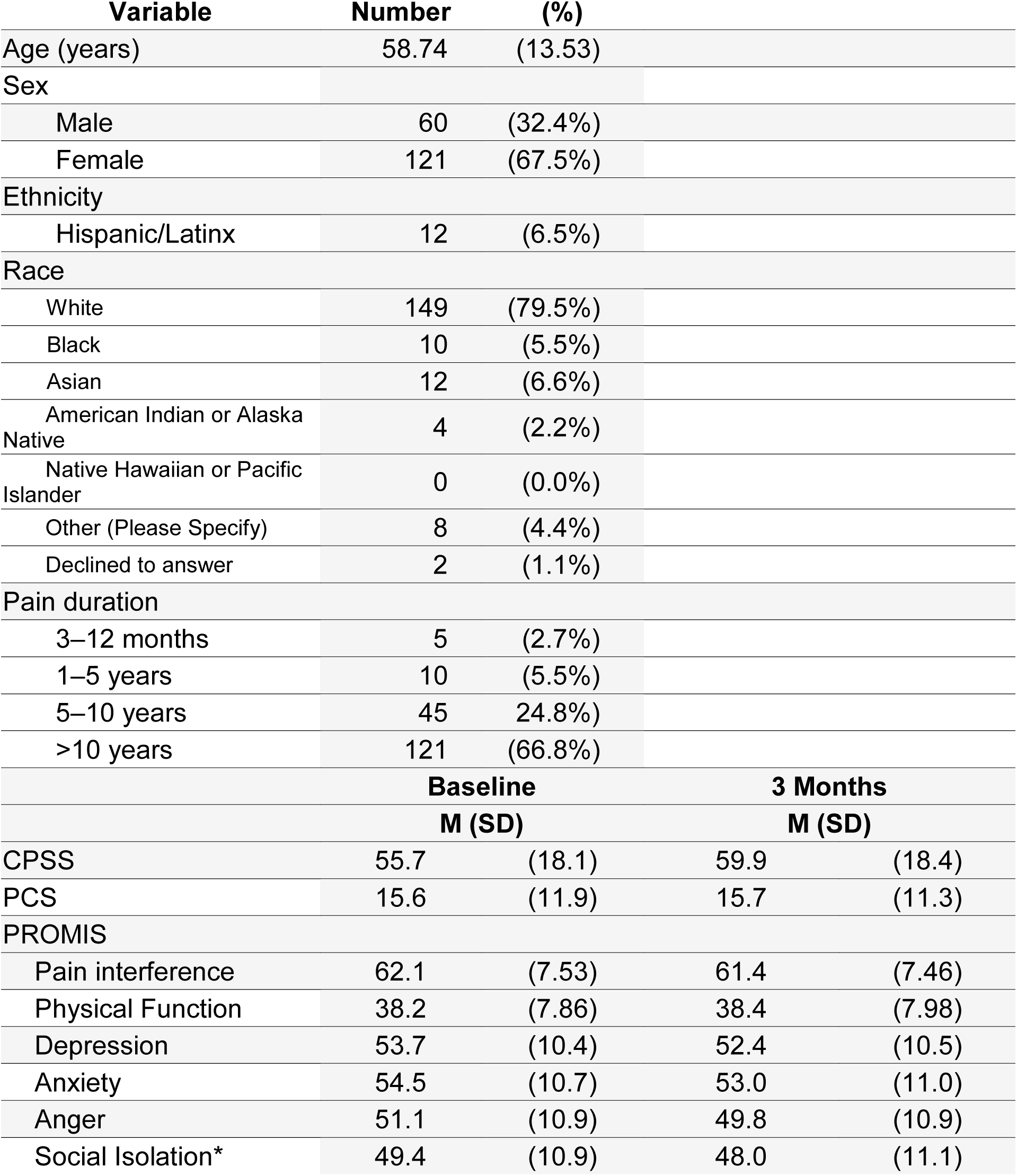
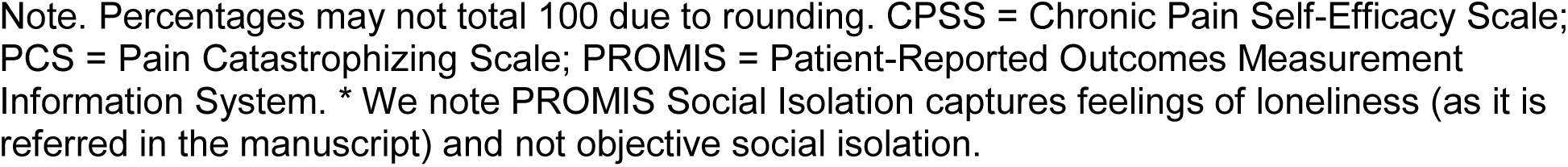
Demographic and Clinical Characteristics (N=181)

| Variable | Number | (%) |  |  |
| --- | --- | --- | --- | --- |
| Age (years) | 58.74 | (13.53) |  |  |
| Sex |  |  |  |  |
| Male | 60 | (32.4%) |  |  |
| Female | 121 | (67.5%) |  |  |
| Ethnicity |  |  |  |  |
| Hispanic/Latinx | 12 | (6.5%) |  |  |
| Race |  |  |  |  |
| White | 149 | (79.5%) |  |  |
| Black | 10 | (5.5%) |  |  |
| Asian | 12 | (6.6%) |  |  |
| American Indian or Alaska Native | 4 | (2.2%) |  |  |
| Native Hawaiian or Pacific Islander | 0 | (0.0%) |  |  |
| Other (Please Specify) | 8 | (4.4%) |  |  |
| Declined to answer | 2 | (1.1%) |  |  |
| Pain duration |  |  |  |  |
| 3–12 months | 5 | (2.7%) |  |  |
| 1–5 years | 10 | (5.5%) |  |  |
| 5–10 years | 45 | 24.8%) |  |  |
| >10 years | 121 | (66.8%) |  |  |
|  | **Baseline** |  | **3 Months** |  |
|  | **M (SD)** |  | **M (SD)** |  |
| CPSS | 55.7 | (18.1) | 59.9 | (18.4) |
| PCS | 15.6 | (11.9) | 15.7 | (11.3) |
| PROMIS |  |  |  |  |
| Pain interference | 62.1 | (7.53) | 61.4 | (7.46) |
| Physical Function | 38.2 | (7.86) | 38.4 | (7.98) |
| Depression | 53.7 | (10.4) | 52.4 | (10.5) |
| Anxiety | 54.5 | (10.7) | 53.0 | (11.0) |
| Anger | 51.1 | (10.9) | 49.8 | (10.9) |
| Social Isolation* | 49.4 | (10.9) | 48.0 | (11.1) |

Note. Percentages may not total 100 due to rounding. CPSS = Chronic Pain Self-Efficacy Scale; PCS = Pain Catastrophizing Scale; PROMIS = Patient-Reported Outcomes Measurement Information System. \* We note PROMIS Social Isolation captures feelings of loneliness (as it is referred in the manuscript) and not objective social isolation.

### Questionnaires

#### Pain Catastrophizing Scale (PCS)

The 13-item PCS assesses pain-related catastrophic thinking on a 5-point Likert scale (0 = “Not at all” to 4 = “All the time”; Cronbach’s α = .94). Total scores range from 0–52, with higher scores indicating greater catastrophizing^7^.

#### Chronic Pain Self-Efficacy Scale (CPSS)

The 22-item CPSS measures confidence in managing pain, daily activities, and pain-related symptoms, rated on a scale from 10 (“Very uncertain”) to 100 (“Very certain”); Cronbach’s α = .89. Scores reflect the average across items (range: 10–100), with higher scores indicating greater pain self-efficacy.

#### Patient-Reported Outcomes Measurement Information System (PROMIS) Measures

PROMIS short forms assessed pain interference (8a), physical function (8b), fatigue (8a), depression (8b), anxiety (8a), anger (8a), and social isolation (8a)^19–20^. Items used 5-point Likert scales. T-scores (M = 50, SD = 10) were calculated using the web-based scoring system (https://www.healthmeasures.net). Higher scores indicate worse outcomes, except for Physical Function, where higher scores indicate better functioning. We also note that PROMIS Social Isolation asks questions about subjective feelings of isolation (i.e., loneliness; ‘I feel that people are around me but not with me’) and not objective isolation. Additionally, it has been previously operationalized as perceived isolation and loneliness^21–22^; therefore, we refer to the item as ‘loneliness’ throughout. Correlation coefficients among PROMIS measures are presented in Table A.

#### Pain Intensity

Pain intensity was measured using one item from the Graded Chronic Pain Scale–Revised (GCPS-R), assessing average pain over the past 7 days (0 = no pain, 10 = worst imaginable pain)^23^.

### Analysis Plan

The primary analyses used hierarchical multiple regression models^24^ to examine whether PCS, CPSS, and their interaction predicted 3-month outcomes and 3-month outcome change scores. Age and sex were included as covariates in all models. Continuous predictor variables (CPSS and PCS) were standardized by subtracting the sample mean and dividing by the sample standard deviation (z = [X − M] / SD). Interaction terms (CPSS × PCS) were computed using these standardized scores to reduce multicollinearity and facilitate interpretation of main and interaction effects in the regression models predicting physical function and psychosocial outcomes.

Effect sizes (R^2^) were calculated to quantify the proportion of variance in the outcomes explained by the predictors. The change in R^2^ (ΔR^2^) was reported to indicate the additional variance explained by the interaction term beyond the main effects (0.02 = small, 0.13 = medium, and 0.26 = large)^9^. False discovery rate (FDR)–adjusted p-values were applied to control for Type I errors due to multiple comparisons. All analyses were conducted in R^25^ (version 4.3.1).

### Baseline Coping Predicting Physical and Psychosocial Outcomes at 3 Months

Hierarchical regression models predicted 3-month outcomes from age, sex (female = 1), PCS, and CPSS. The PCS × CPSS interaction was added in Step 2 to determine whether the interaction explained additional variance beyond the main effects. Separate models were conducted for pain intensity, pain interference, physical function, depression, anxiety, anger, and loneliness.

### Baseline Coping Predicting Change in Physical and Psychosocial Outcomes Over 3 Months

Baseline-adjusted multiple regression models predicted 3-month change scores from age, sex (female = 1), PCS, CPSS, and PCS × CPSS interaction. Each model included the baseline level of the outcome to assess changes over 3 months. Separate models were conducted for pain intensity, pain interference, physical function, depression, anxiety, anger, and loneliness.

### Power Analysis

Using G*Power^26^ 3.1.9.7, the required sample size for detecting a medium effect (f^2^ = 0.15) with 5 predictors, α = .05, and power = 0.99 was N = 125. The final sample (N = 181) exceeded this threshold.

### Missing Data

Baseline data were complete. At 3-month follow-up, all measures were complete except for pain intensity (missing in 5.5% of cases). Given that missingness was minimal (≤5.5%), pairwise deletion was used to maximize the sample size.

## Results

### Participants represented long-duration chronic pain with moderate psychosocial impacts and homogenous demographics

Demographics are summarized in Table 1. Participants were 181 adults receiving care for chronic pain who completed baseline and 3-month follow-up assessments online. The mean age of the sample was 58.74 years (SD = 13.53). Most participants identified as female (66.7%) and White (79.5%), with additional representation from Black (5.5%), Asian (6.6%), and other racial groups (7.7%), which included American Indian or Alaska Native and Native Hawaiian or Pacific Islander, and “Other”, which, when selected, prompted the patient to “please specify” with a text box. A minority of participants identified as Hispanic/Latinx (6.5%). Chronic pain duration was typically long-standing, with 66.8% reporting pain lasting more than 10 years. Reported pain locations were primarily low back (40.2%), neck (34.8%), and lower extremities (25.0%).

On average, our participants reported moderate pain self-efficacy (CPSS: M_Baseline_ = 55.7; M_3-months_ = 59.9) and moderate pain catastrophizing (PCS: M_Baseline_ = 15.6; M_3-months_ = 15.7). Regarding PROMIS outcomes, participants reported moderate pain interference (M_Baseline_ = 62.1; M_3-months_ = 61.4) and low physical function (M_Baseline_ = 38.2; M_3-months_ = 38.4). Participants reported average levels of depression, anxiety, anger, and loneliness at both time points (M_Baseline_ = 49.4 ∼ 54.5; M_3-months_ = 48.0 ∼ 53.0).

**Hypothesis 1:** The interaction between pain catastrophizing and pain self-efficacy will significantly predict physical and psychosocial outcomes at 3 months and improve prediction beyond main effects.

Models predicting physical outcomes are presented in Table 2. Including an interaction term (PCS x CPSS) improved our model fit only for physical function (Δ*R*^2^ = 0.02, *p* = .022). Pain self-efficacy was positively associated with physical function at 3 months (*β* = 0.646. p < .001), but this positive association weakened at high levels of PCS (Figure 1). PCS scores were positively associated with pain intensity (*β* = 0.212, *p* = .009) and pain interference (*β* = 0.164, *p* = .030), whereas CPSS scores were negatively associated with pain intensity (*β* = −0.330, *p* < .001) and pain interference (*β* = −0.450, *p* < .001). Notably, the effect sizes of our predictive model ranged from medium (pain intensity *R*^2^*=* .220) to large (*R*^2^ *_Range_=* .309-.458).

**Figure 1.**
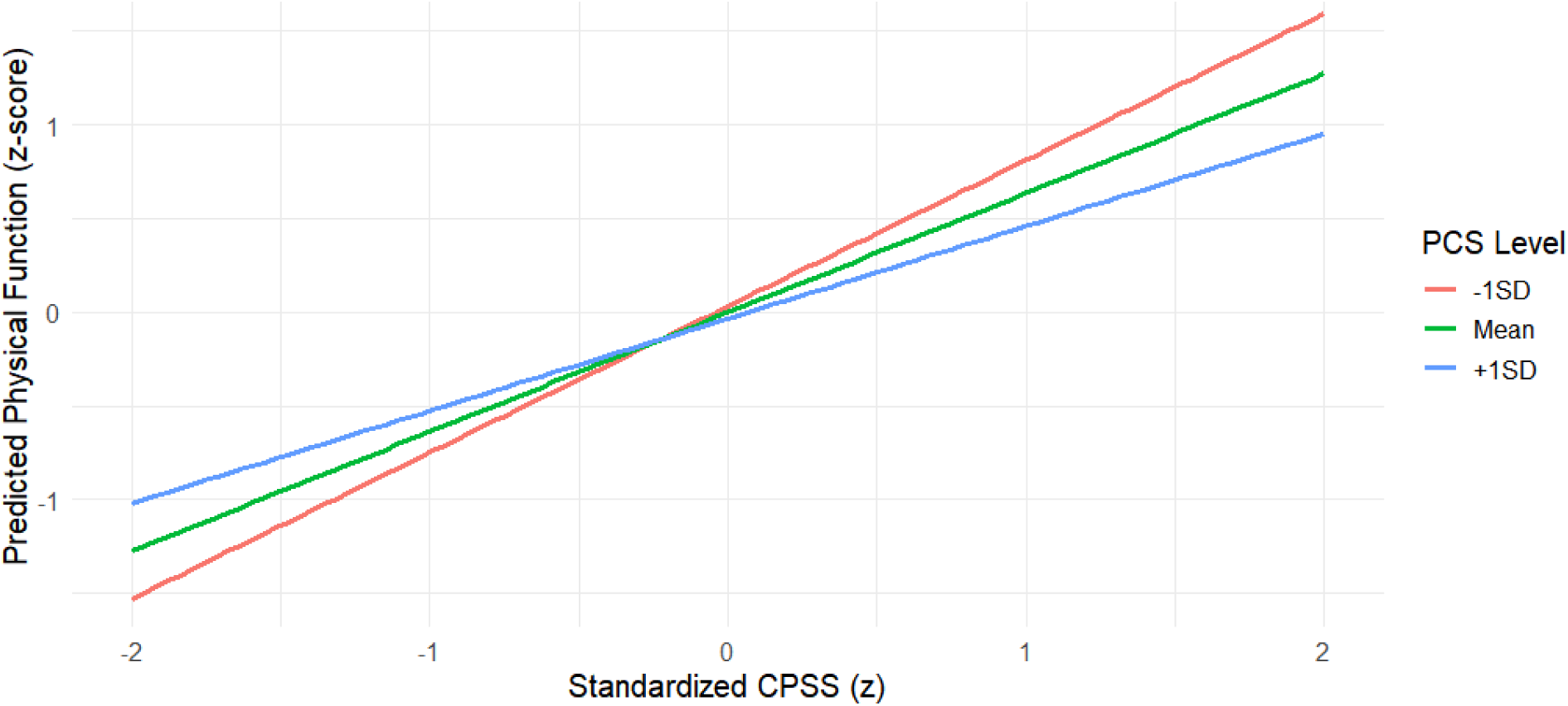
Interaction between CPSS and PCS. The predictive effect of CPSS on physical function at 3 months decreases as PCS scores increase.

**Table 2.** Results of stepwise regression models examining whether the interaction between PCS and CPSS significantly improves the prediction of pain, pain interference, and physical function at 3-month follow-up.

| Average Pain Intensity (N = 171) |  |  |  |  |  |  |  |
| --- | --- | --- | --- | --- | --- | --- | --- |
| Step 1 | *B* | *SE* | $\beta$ | *p* | *F* | *P* | $R^2$ |
| Age | 0.014 | 0.011 | 0.087 | .278 | 11.70 | < .001 | .220 |
| Female | 0.244 | 0.329 | 0.053 | .461 |  |  |  |
| **PCS** | **0.040** | **0.014** | **0.212** | **.009** |  |  |  |
| **CPSS** | **-0.038** | **0.009** | **-0.330** | **< .001** |  |  |  |
| Step 2 | | | | | | | $\Delta R^2$ |
| PCS x CPSS | 0.001 | .001 | .240 | .376 |  |  | .008 |
| Pain Interference (N = 181) |  |  |  |  |  |  |  |
| Step 1 | *B* | *SE* | $\beta$ | *p* | *F* | *P* | $R^2$ |
| Age | 0.059 | 0.036 | 0.108 | .123 | 19.67 | < .001 | .309 |
| Female | 1.530 | 1.035 | 0.096 | .141 |  |  |  |
| **PCS** | **0.108** | **0.046** | **0.164** | **.030** |  |  |  |
| **CPSS** | **-0.182** | **0.028** | **-0.450** | **< .001** |  |  |  |
| Step 2 | | | | | | | $\Delta R^2$ |
| PCS x CPSS | 0.003 | 0.002 | 0.244 | .199 |  |  | .008 |
| Physical Function (N = 181) |  |  |  |  |  |  |  |
| Step 1 | *B* | *SE* | *B* | *p* | *F* | *P* | *R*<sup>2</sup> |
| Age | -0.079 | 0.034 | -0.134 | .026 | 37.24 | < .001 | .458 |
| Female | -2.326 | 0.973 | -0.136 | .026 |  |  |  |
| PCS | 0.000 | 0.043 | -0.001 | .992 |  |  |  |
| **CPSS** | **0.281** | **0.027** | **0.646** | **< .001** |  |  |  |
| Step 2 | | | | | | | $\Delta R^2$ |
| **PCS x CPSS** | **-0.005** | **0.002** | **-0.380** | **.022** |  |  | **.020** |

Note: FDR adjusted p values were noted. Values in bold indicate statistical significance. Pain intensity (N=171) differs due to survey completion.

Models predicting psychosocial outcomes are presented in Table 3. No significant interaction effects were observed for any psychosocial outcomes at 3-months follow-up. Significant main effects emerged such that PCS positively predicted depression (*β* = 0.512), anxiety (*β* = 0.535), anger (*β* = 0.451), and loneliness (*β* = 0.307; all *p’s* < .001). Higher CPSS also predicted lower loneliness (*β* = − 0.247, *p* = .001) but did not for our other psychosocial measures (*p’s* >.05). Notably, model effect sizes ranged from medium (*R*^2^*=* .234 for anger) to large (*R^2^_range_=* .264-.359 for loneliness, anxiety, and depression).

**Table 3.** Results of stepwise regression models examining whether the interaction between PCS and CPSS significantly improves the prediction of depression, anxiety, anger, and social isolation at 3-month follow-up.

| Depression (N = 181) |  |  |  |  |  |  |  |
| --- | --- | --- | --- | --- | --- | --- | --- |
| Step 1 | *B* | *SE* | $\beta$ | *p* | *F* | *P* | *R*<sup>2</sup> |
| Age | -0.033 | 0.048 | -0.042 | .499 | 24.02 | < .001 | .353 |
| Female | 1.996 | 1.400 | 0.090 | .195 |  |  |  |
| **PCS** | **0.474** | **0.062** | **0.512** | **< .001** |  |  |  |
| CPSS | -0.073 | 0.038 | -0.130 | .090 |  |  |  |
| Step 2 | | | | | | | $\Delta R^2$ |
| PCS x CPSS | 0.001 | 0.003 | 0.069 | .678 |  |  | .001 |
| Anxiety (N = 181) |  |  |  |  |  |  |  |
| Step 1 | *B* | *SE* | $\beta$ | *p* | *F* | *P* | *R*<sup>2</sup> |
| Age | -0.087 | 0.050 | -0.107 | .145 | 24.69 | < .001 | .359 |
| Female | 1.752 | 1.465 | 0.075 | .243 |  |  |  |
| **PCS** | **0.520** | **0.064** | **0.535** | **< .001** |  |  |  |
| CPSS | -0.046 | 0.039 | -0.078 | .243 |  |  |  |
| Step 2 | | | | | | | $\Delta R^2$ |
| PCS x CPSS | 0.001 | 0.003 | 0.047 | .774 |  |  | .000 |
| Anger (N = 181) |  |  |  |  |  |  |  |
| Step 1 | *B* | *SE* | $\beta$ | *p* | *F* | *P* | *R*<sup>2</sup> |
| Age | -0.080 | 0.055 | -0.100 | .243 | 13.43 | < .001 | .234 |
| Female | -0.275 | 1.594 | -0.012 | .863 |  |  |  |
| **PCS** | **0.437** | **0.070** | **0.451** | **< .001** |  |  |  |
| CPSS | -0.020 | 0.043 | -0.034 | .796 |  |  |  |
| Step 2 | | | | | | | $\Delta R^2$ |
| PCS x CPSS | -0.005 | 0.002 | -0.016 | .928 |  |  | .000 |
| Loneliness (N = 181) |  |  |  |  |  |  |  |
| Step 1 | *B* | *SE* | $\beta$ | *p* | *F* | *p* | *R*<sup>2</sup> |
| Age | -0.090 | 0.055 | -0.109 | .106 | 15.75 | < .001 | .264 |
| Female | 2.699 | 1.605 | 0.113 | .106 |  |  |  |
| **PCS** | **0.305** | **0.071** | **0.307** | **< .001** |  |  |  |
| **CPSS** | **-0.150** | **0.043** | **-0.247** | **.001** |  |  |  |
| Step 2 | | | | | | | $\Delta R^2$ |
| PCS x CPSS | 0.004 | 0.003 | 0.192 | .329 |  |  | .005 |

Note: FDR adjusted $p$ values were noted. Values in bold indicate statistical significance.

**Hypothesis 2:** The interaction between pain catastrophizing and pain self-efficacy will significantly predict changes in physical and psychosocial outcomes over time.

Models predicting changes in physical outcomes are presented in Table 4. We observed a significant interaction only for changes in physical function (*p* = .025); however, the FDR-corrected p-value was marginal (*p* = .059). When controlling for baseline levels, neither PCS nor CPSS predicted changes in physical health outcomes. Model effect sizes, including the baseline scores, were large (*R*^2^ = .527 ∼ .798).

**Table 4.**
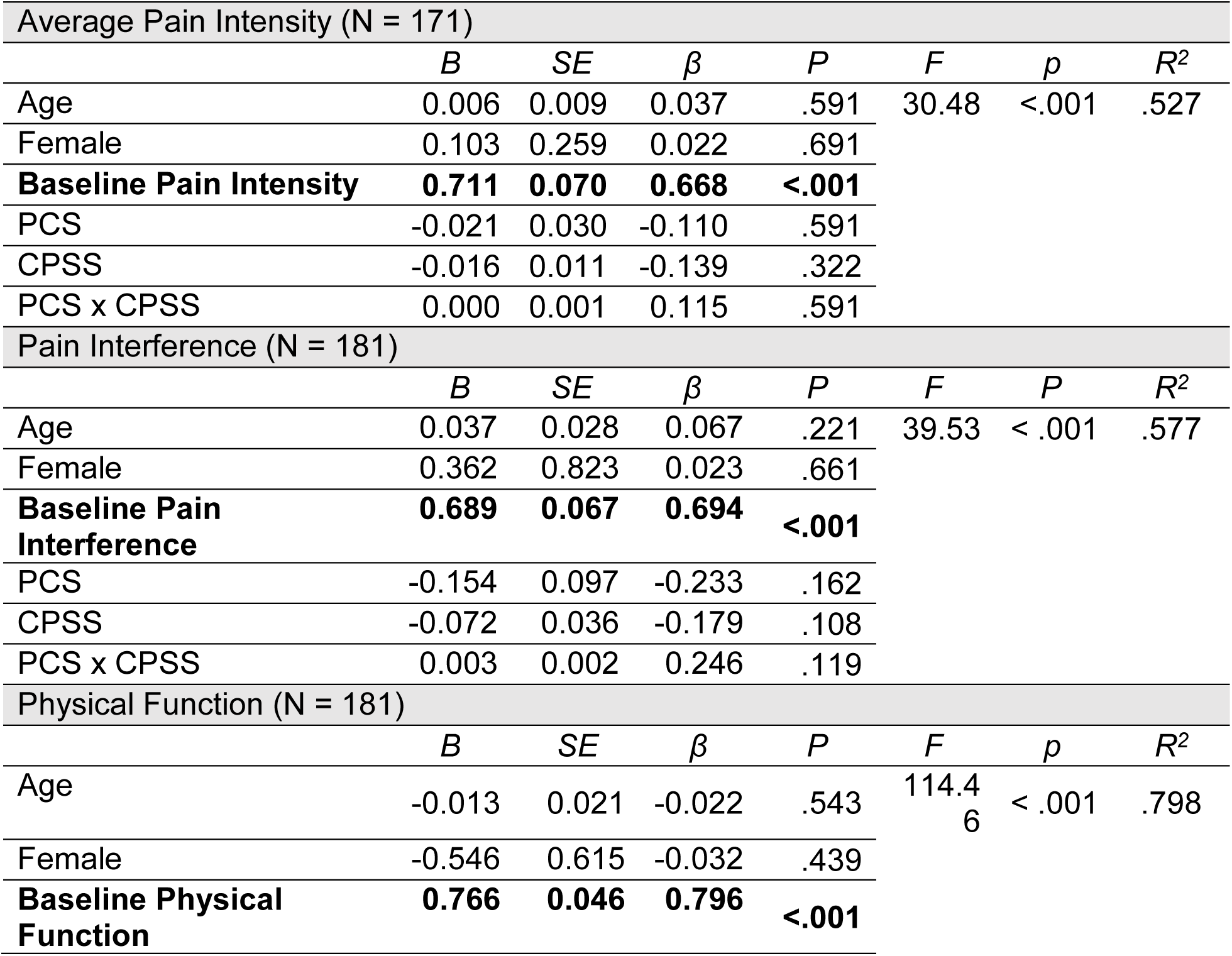
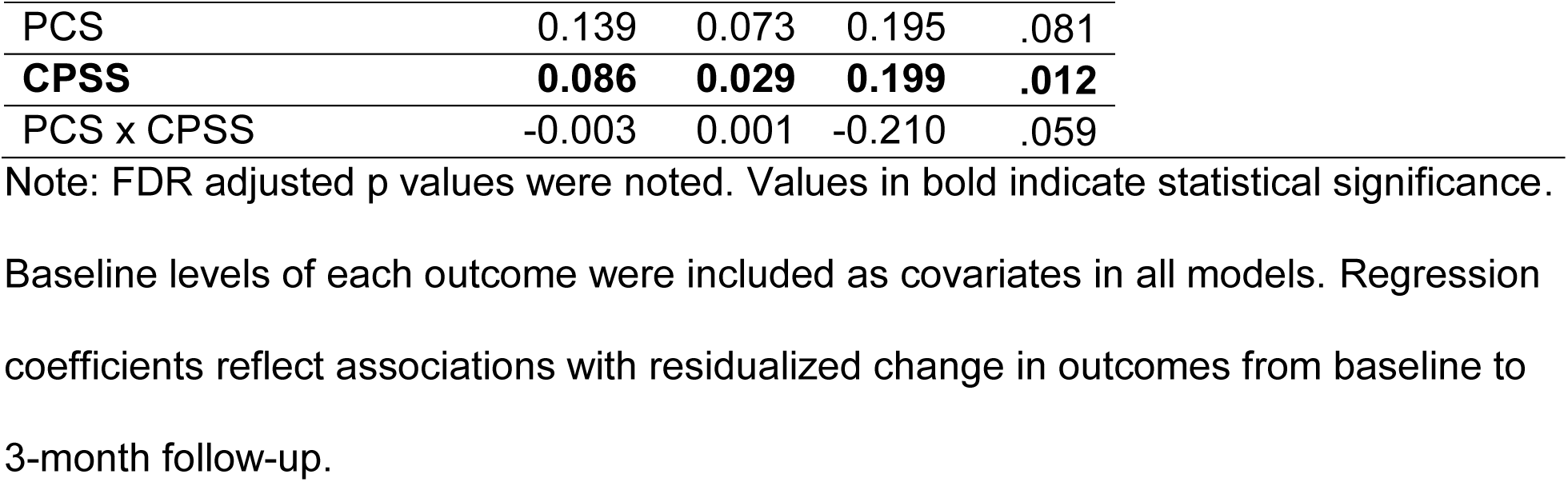
Results of multiple regression models predicting physical health outcomes at 3-month follow-up, adjusted for baseline outcomes.

| Average Pain Intensity (N = 171) |  |  |  |  |  |  |  |
| --- | --- | --- | --- | --- | --- | --- | --- |
| | *B* | *SE* | $\beta$ | *P* | *F* | *p* | *R*<sup>2</sup> |
| Age | 0.006 | 0.009 | 0.037 | .591 | 30.48 | <.001 | .527 |
| Female | 0.103 | 0.259 | 0.022 | .691 |  |  |  |
| **Baseline Pain Intensity** | **0.711** | **0.070** | **0.668** | **<.001** |  |  |  |
| PCS | -0.021 | 0.030 | -0.110 | .591 |  |  |  |
| CPSS | -0.016 | 0.011 | -0.139 | .322 |  |  |  |
| PCS x CPSS | 0.000 | 0.001 | 0.115 | .591 |  |  |  |
| Pain Interference (N = 181) |  |  |  |  |  |  |  |
| | *B* | *SE* | $\beta$ | *P* | *F* | *P* | *R*<sup>2</sup> |
| Age | 0.037 | 0.028 | 0.067 | .221 | 39.53 | < .001 | .577 |
| Female | 0.362 | 0.823 | 0.023 | .661 |  |  |  |
| **Baseline Pain Interference** | **0.689** | **0.067** | **0.694** | **<.001** |  |  |  |
| PCS | -0.154 | 0.097 | -0.233 | .162 |  |  |  |
| CPSS | -0.072 | 0.036 | -0.179 | .108 |  |  |  |
| PCS x CPSS | 0.003 | 0.002 | 0.246 | .119 |  |  |  |
| Physical Function (N = 181) |  |  |  |  |  |  |  |
| | *B* | *SE* | $\beta$ | *P* | *F* | *p* | *R*<sup>2</sup> |
| Age | -0.013 | 0.021 | -0.022 | .543 | 114.46 | < .001 | .798 |
| Female | -0.546 | 0.615 | -0.032 | .439 |  |  |  |
| **Baseline Physical Function** | **0.766** | **0.046** | **0.796** | **<.001** |  |  |  |

|  |  |  |  |  |
| --- | --- | --- | --- | --- |
| PCS | 0.139 | 0.073 | 0.195 | .081 |
| **CPSS** | **0.086** | **0.029** | **0.199** | **.012** |
| PCS x CPSS | -0.003 | 0.001 | -0.210 | .059 |

Note: FDR adjusted p values were noted. Values in bold indicate statistical significance.

Baseline levels of each outcome were included as covariates in all models. Regression coefficients reflect associations with residualized change in outcomes from baseline to 3-month follow-up.

Models predicting changes in psychosocial outcomes are presented in Table 5. When controlling for baseline scores, PCS, CPSS, and their interaction predicted changes in loneliness only. Higher CPSS scores were associated with reductions in loneliness over time (*β* = −0.177, *p* = .012). However, this negative association weakened at higher levels of PCS (Figure 2). Model effect sizes, including the baseline scores, were large (*R*^2^ = .544 ∼ .753).

**Figure 2.**
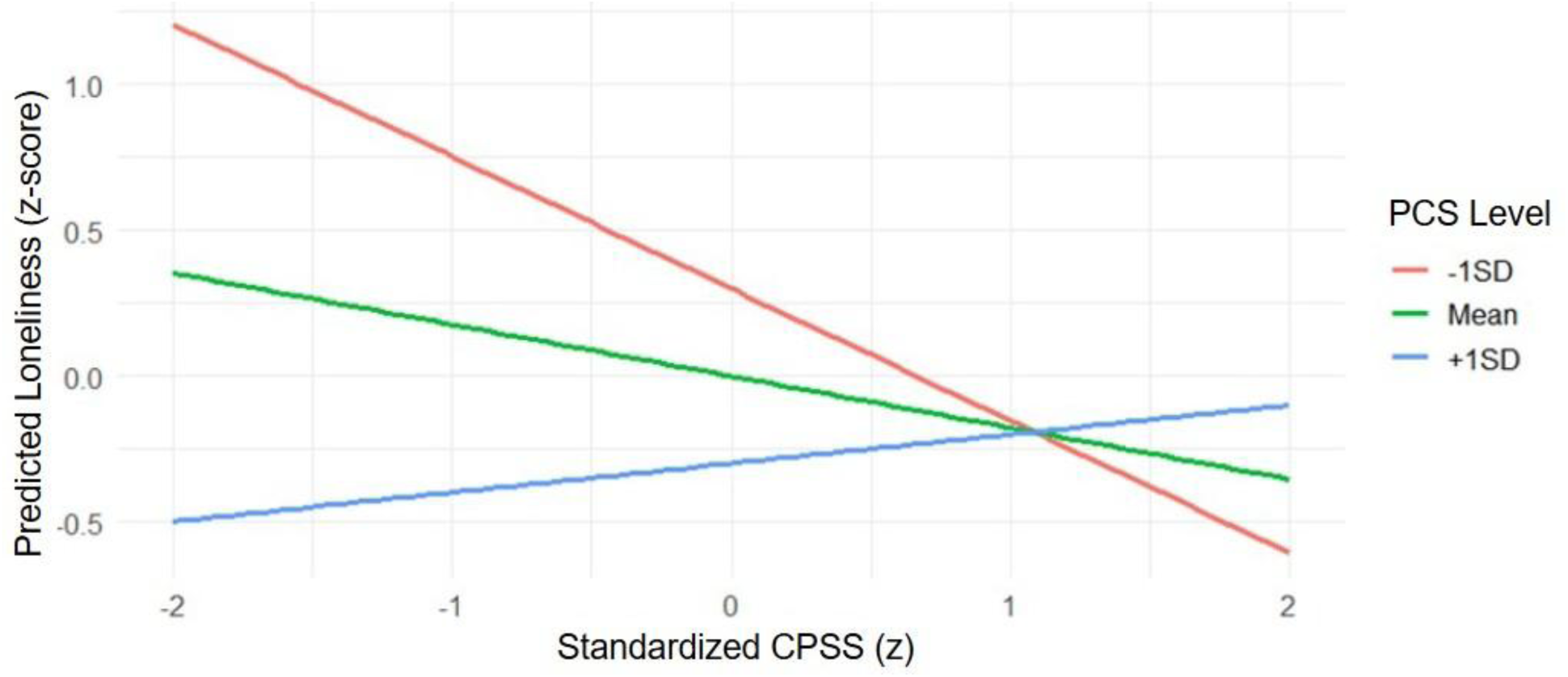
Significant Interaction between CPSS and PCS. The predictive effect of CPSS on the 3-month changes in loneliness decreases as PCS scores increase.

**Table 5.**
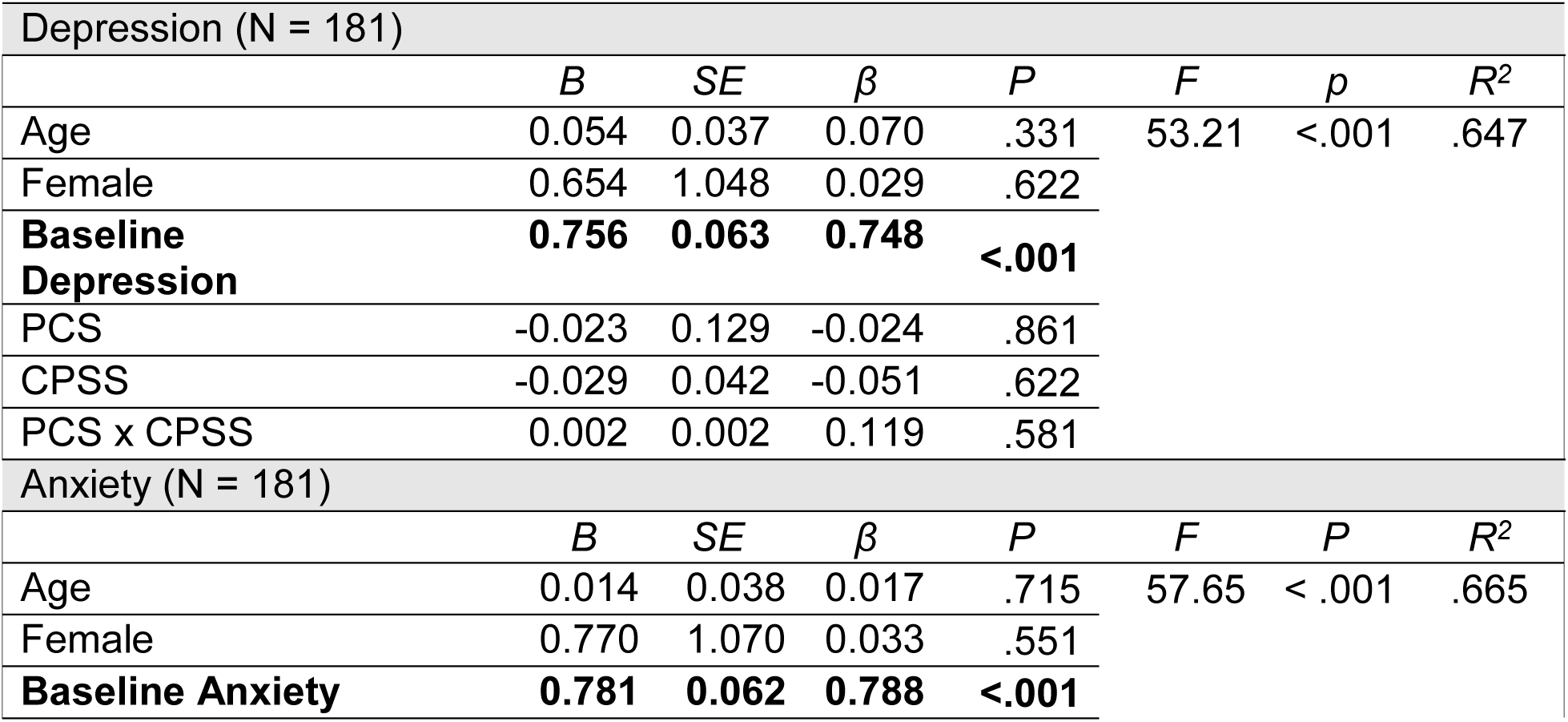
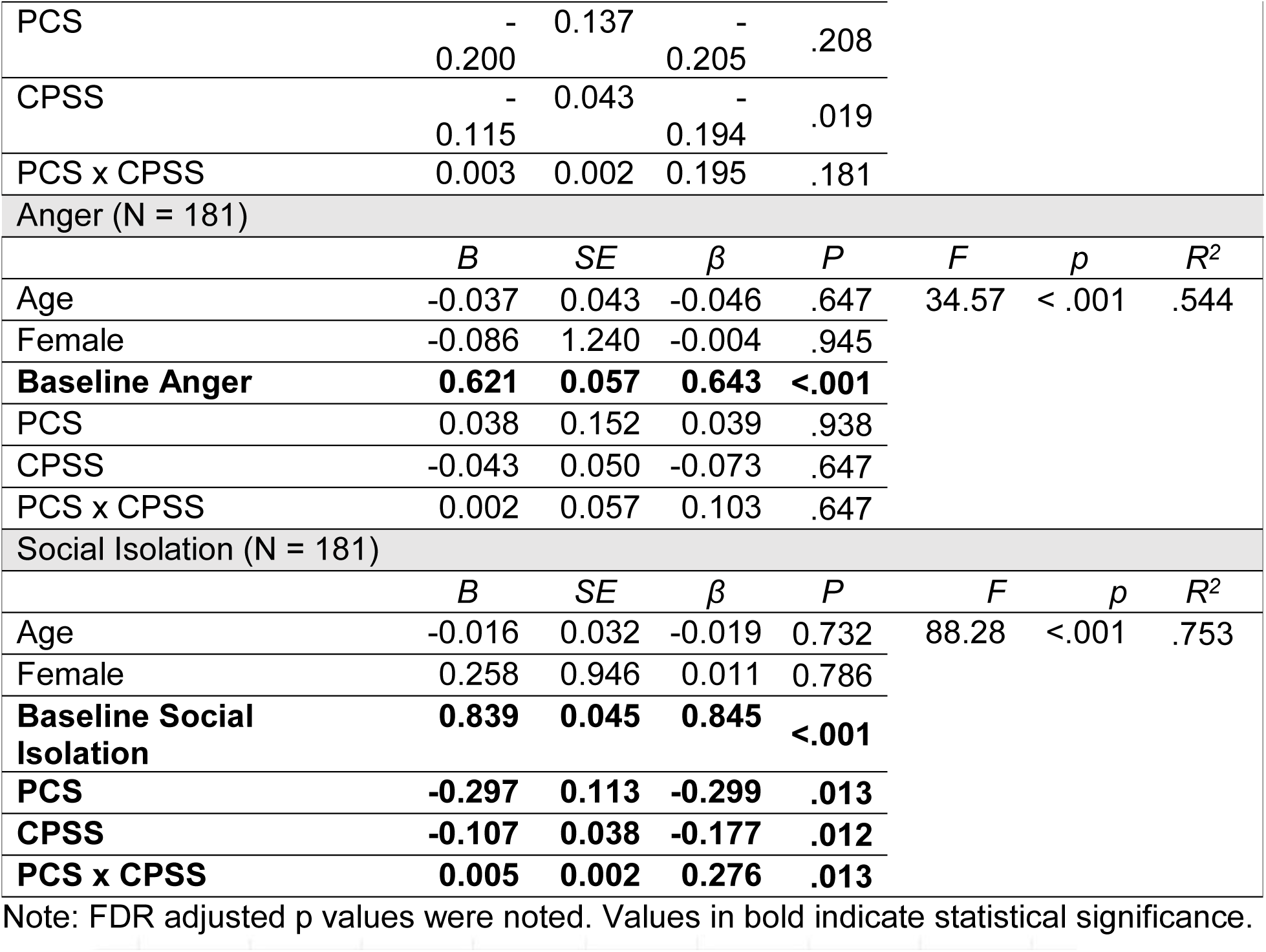
Results of multiple regression models predicting psychosocial health outcomes at 3-month follow-up when controlling for the baseline outcomes.

### Correlational Analysis

To characterize the relationships among key chronic pain outcomes across time, we computed Pearson correlations among all baseline and 3-month follow-up variables (Table 2). The upper-right triangle of the table presents baseline correlations, the middle-left triangle presents follow-up correlations, and the lower-left triangle provides cross-time correlations between baseline and follow-up scores.

**Table A.** Pearson correlations among outcome variables at baseline and 3-month follow-up. PCS and CPSS were moderately inversely correlated (r = –0.41, p < .001; Supplemental Table 2)

|  | 1. | 2. | 3. | 4. | 5. | 6. | 7. |
| --- | --- | --- | --- | --- | --- | --- | --- |
| 1. Average Pain Intensity |  | .666 | -.528 | .350 | .319 | .198 | .311 |
| 2. Pain Interference | .702 |  | -.661 | .472 | .374 | .281 | .508 |
| 3. Physical Function | -.591 | -.699 |  | -.310 | -.218 | -.116 | -.306 |
| 4. Depression | .393 | .497 | -.321 |  | .802 | .612 | .744 |
| 5. Anxiety | .337 | .384 | -.242 | .782 |  | .635 | .626 |
| 6. Anger | .312 | .407 | -.232 | .695 | .672 |  | .573 |
| 7. Social Isolation | .269 | .470 | -.327 | .710 | .660 | .692 |  |
| 1. Average Pain Intensity | .720 |  |  |  |  |  |  |
| 2. Pain Interference | .562 | .749 |  |  |  |  |  |
| 3. Physical Function | -.518 | -.633 | .886 |  |  |  |  |
| 4. Depression | .337 | .448 | -.262 | .796 |  |  |  |
| 5. Anxiety | .330 | .380 | -.211 | .683 | .805 |  |  |
| 6. Anger | .213 | .376 | -.160 | .585 | .629 | .722 |  |
| 7. Social Isolation | .261 | .478 | -.336 | .662 | .554 | .445 | .860 |

Upper right (light blue) triangle shows baseline correlations; middle left (green) triangle shows 3-month follow-up correlations; lower left (light orange) triangle shows correlations between baseline and 3-month follow-up. All $p$ 's $< .001$ . All outcomes had $n = 181$ , except average pain intensity ( $n = 171$ ).

## Discussion

This study examined whether pain catastrophizing and pain self-efficacy interact to predict health outcomes for patients living with chronic pain. We identified significant interactions between pain-related catastrophizing and self-efficacy that provided greater nuance to psychosocial and physical functioning in chronic pain. First, the inclusion of our interaction term improved the prediction of physical function at three months; however, this effect was attenuated after false discovery rate (FDR) correction. The opposite emerged for loneliness, in which interactions did not predict loneliness at three months but did predict changes in loneliness over three months. Across outcomes, we replicated main effects of both pain catastrophizing (PCS) and pain self-efficacy (CPSS) for pain intensity, pain interference, and loneliness, with greater catastrophizing and lower self-efficacy associated with poorer pain-related outcomes^9,12^. For psychological distress (i.e., depression, anxiety, and anger), PCS, as expected was consistently positively associated; however, CPSS only associated with anxiety and only after controlling for baseline levels.

Our findings align with broader evidence indicating that higher baseline pain self-efficacy is associated with better physical function and has been linked to greater improvement following multidisciplinary treatment^13^. Considering physical health outcomes, physical function and pain interference were moderately related but represented distinct dimensions of chronic pain outcomes. Physical function reflects perceived ability to perform instrumental activities of daily living (e.g., vacuuming or yard work) and movement-related activities (e.g., walking for 15 minutes)^27^, whereas pain interference reflects perceived limitations in engagement with activities due to pain^28^. Importantly, pain self-efficacy is considered a modifiable construct and will be an important target to consider for chronic pain interventions^29–30^. Consistent with this distinction, pain self-efficacy robustly predicted physical functioning in the present study of patients treated at a tertiary pain clinic and both pain catastrophizing and pain self-efficacy demonstrated significant main effects for pain interference, but neither construct predicted changes in pain interference over time. Notably, behavioral interventions appear to exert stronger effects on reducing pain catastrophizing than enhancing pain self-efficacy, but this may be due in part to the imbalance in RCT representation between the two constructs. Given the interactive properties with pain catastrophizing and pain self-efficacy observed in the present study, collecting both measures seems important for future RCTs for chronic pain and achieving low levels of pain catastrophizing may be particularly important to reap the benefits of self-efficacy. At the same time, an important next step for the field is to develop strategies that more effectively enhance pain self-efficacy^29^.

This study also provides initial evidence that adaptive and maladaptive pain cognitions may interact in predicting loneliness among patients with chronic pain. After adjusting for baseline levels of loneliness, the interaction between pain self-efficacy and pain catastrophizing significantly predicted loneliness at three months. Specifically, higher baseline self-efficacy was associated with lower subsequent loneliness; however, this protective effect was attenuated at higher levels of catastrophizing. In contrast, when baseline loneliness was not accounted for, pain self-efficacy and pain catastrophizing independently predicted loneliness, suggesting that their interactive effects may be most evident when modeling change over time. These findings extend prior work indicating that loneliness is often resistant to improvement with treatment in chronic pain populations^31,32^. Rather than reflecting uniform stability or change, our results suggest that loneliness may be differentially influenced by cognitive factors. This pattern highlights specific cognitive targets (i.e., PCS and CPSS) that may be especially relevant for addressing social dysfunction and its downstream consequences, such as poor reintegration and loss of work^33,34^. Clinically, this distinction is meaningful, as higher levels of loneliness have been associated with reduced treatment engagement and poorer treatment response^35^.

Loneliness was weakly correlated with physical function (r = −0.327 at 3 months), suggesting physical ability is less dependent on social connections. In contrast, loneliness was strongly associated with depression (r = 0.710 at 3 months), consistent with prior literature linking loneliness and depression in chronic pain^36^ and showing loneliness’ impact on pain interference is partially mediated by depression^32^. Notably, the interaction between pain self-efficacy and catastrophizing did not extend to depression, anxiety, or anger suggesting the combined effects of self-efficacy and catastrophizing may be more relevant for loneliness than for other affective domains. Previously, it has been suggested that loneliness and social expectations are relatively inflexible (i.e., hard to change or treat) in patients with depression^37^. Although we see loneliness is still correlated with depression here, our data suggests that patients with chronic pain still exhibit fluctuations in their levels of loneliness based on their levels of catastrophizing (up-regulate) and self-efficacy (down-regulate) independent of depression. However, further investigation into whether depression mediates any of the effects we observed here from self-efficacy and catastrophizing on loneliness are needed.

Several limitations should be considered. First, the follow-up period was relatively short (3 months), and outcomes relied exclusively on self-report measures. Future studies with longer follow-up periods and objective assessments of physical function (e.g., actigraphy or wearable devices) are needed to confirm and extend these findings. Additionally, participants were recruited from a tertiary pain clinic research registry, ensuring verification of chronic pain status but limiting interpretation of treatment effects, as participants varied in whether they were actively receiving or had completed treatment and had limited variability in terms of demographic representation. Future research should examine long-term trajectories of pain catastrophizing and self-efficacy and develop interventions that concurrently target both constructs to optimize functional and psychosocial outcomes. Further, research should incorporate patient-engaged frameworks^38^ to ensure trials are representative^39^ to improve external validity^40^ and scientific rigor and reduce burden and ability to access interventions^41^. Future research should address two key questions: (1) which combinations of multidisciplinary approaches best optimize functional recovery, and (2) whether tailoring treatment to individuals’ cognitive profiles will maximize functional improvement. Individualized strategies may be informed by targeting the four sources of self-efficacy: (1) mastery experiences, (2) vicarious learning, (3) verbal persuasion, and (4) physiological and emotional states.

## Conclusion

Interactions between pain self-efficacy and pain catastrophizing should be considered when predicting physical function and loneliness in chronic pain. Historically, pain psychology research has focused on the prognostic ability of negative pain cognitions and less on protective cognitions. Despite the likelihood that individual differences in pain outcomes may depend on the relative use of maladaptive and adaptive pain cognitions, even fewer studies have examined the prognostic ability of their interaction,. Although enhancing adaptive and lowering maladaptive pain cognitions are independently well-established strategies for improving clinical outcomes, the present study highlights the importance of considering their interaction in predicting physical and social functioning. These cognitive and modifiable prognostic factors may represent key treatment targets for optimizing function in patients with chronic pain.

## Acknowledgments

We acknowledge funding from NIH T32 DA035165 (TD, SK), and the Redlich Fund, courtesy of Dr. Sean Mackey. (ER)

## Disclosures

The author(s) report no conflicts of interest in this work.

## Data Availability

All data produced in the present study are available upon reasonable request to the authors.

## Notes

### Competing Interest Statement

The authors have declared no competing interest.

### Author Declarations

Institutional Review Board of Stanford University gave ethical approval for this work.

